# Longitudinal Patterns of Digital Parenting Restrictions and Adolescent Screen Use: Insights from the Adolescent Brain Cognitive Development (ABCD) Study

**DOI:** 10.1101/2025.11.23.25340847

**Authors:** Daniel Lopez, Arturo Lopez-Flores, Gloria Ruiz-Orozco, Bonnie Nagel

## Abstract

**Objective:** Digital parenting restrictions are widely used to manage adolescent screen use, yet it is unclear how effective these strategies remain as children age and gain autonomy. This study examined longitudinal patterns of digital parenting restrictions and their relationship to adolescent screen time using self-reported and passive measures.

**Method:** Data came from Years 3–5 of the Adolescent Brain Cognitive Development (ABCD) Study (n = 8,096). Latent Transition Analysis identified Low, Moderate, and High Restriction profiles from 14 Parent Screen Time Questionnaire items. Linear mixed-effects models tested associations between profiles and screen use outcomes (self-reported screen time, self-reported smartphone use, passive smartphone use from the ABCD-EARS app, and smartphone keystroke activity). Interaction terms examined age effects. Gradient boosting (XGBoost) identified the most influential parenting practices.

**Results:** High and Moderate Restriction profiles were linked to lower screen use across all measures. Compared to the Low Restriction group, High Restriction participants reported 56 fewer daily minutes of screen time (p < 0.0008) and logged 286 fewer daily keystrokes (p < 0.0008). A significant interaction (β = 13.0; 95% CI: 7.9, 18.2) indicated that the effect of high restriction diminished with age. Machine learning highlighted bedroom device access (e.g., screens playing at bedtime, mobile devices in bed) as the strongest predictor of higher screen time.

**Conclusion:** Parenting restrictions reduce adolescent screen time, but their impact declines as adolescents gain independence. Limiting device access in the bedroom may be one of the most effective and lasting strategies for moderating screen use and fostering healthier digital habits.

## Introduction

### Background

The use of digital media devices, including smartphones, computers, and streaming platforms, has become a routine part of daily life for children and adolescents. In the United States, more than half of teenagers aged 12–17 report spending four or more hours per day on screens, with usage increasing steadily as children grow older (Zablotsky, Arockiaraj, Haile, & Ng, 2024). While digital media use offers benefits such as facilitating social connections, supporting educational pursuits, and enabling creative skills like video content production (Blum-Ross & Livingstone, 2018), concerns persist regarding its risks. High screen time has been linked to negative health outcomes, including disrupted sleep patterns, diminished academic performance, and delays in cognitive development (Lissak, 2018; Muppalla, Vuppalapati, Pulliahgaru, Sreenivasulu, & kumar Muppalla, 2023). Although the quality and nature of screen content are crucial, the sheer duration of screen time remains a significant factor influencing adolescent health.

Parents play a central role in regulating children’s screen use. Parenting strategies such as setting time limits, monitoring activities, and encouraging alternative behaviors are critical in shaping digital habits. However, the long-term effectiveness of these practices, especially as children transition into adolescence and gain greater autonomy, remains unclear. Adolescence is a developmental period marked by increasing independence, making it more challenging for parents to enforce restrictions. As children mature, they often gain unsupervised access to digital devices, further complicating parental control.

Research highlights this evolving dynamic. For instance, a meta-analysis examining parental mediation and screen use found that the effectiveness of restrictive parenting strategies declines as children grow older (L. Chen & Shi, 2019). Similarly, Sanders, Parent, Forehand, Sullivan, and Jones (2016) observed that while negative parental attitudes toward technology were linked to lower screen time among younger children, this association diminished during adolescence. This suggests that as children age, developmental changes may outweigh parental attempts at regulating media use.

Despite these insights, important gaps in the literature remain. Existing studies have largely relied on self-reported measures of screen time, which are prone to recall and social desirability biases. For example, recent findings from the Adolescent Brain Cognitive Development (ABCD) Study revealed substantial discrepancies between self-reported smartphone use and passively collected data, with children underreporting their daily smartphone use by more than 2 hours per day (Alexander et al., 2024). Moreover, research examining the impact of digital parenting restrictions over time, particularly using objective, passive assessment methods, is limited. Most available studies have failed to explore how parenting strategies influence adolescent screen use across multiple time points while incorporating passive measurement tools such as smartphone or keystroke data, which can provide novel insights into daily media engagement.

### Study Aims

To address these gaps, the present study examines the longitudinal relationship between digital parenting restrictions and adolescent screen use over three time points, drawing on data from the year-3, year-4, and year-5 visits of the ABCD Study. Unlike previous research, this study integrates both self-reported measures of screen time and passively collected smartphone use data, including screen time duration on Android devices and keystroke activity on both Android and iOS devices. Parenting restrictions were assessed using the Parent Screen Time Questionnaire (PSQ). By identifying distinct parenting profiles based on restriction patterns, this approach allows for a comprehensive examination of all PSQ items simultaneously, rather than selectively focusing on individual items. In addition, machine learning techniques were employed to explore specific parental practices, such as bedroom device access, that may predict changes in adolescent screen use over time.

This research hypothesizes that higher levels of digital parenting restrictions will be associated with lower screen time across both self-reported and passively measured outcomes. It also anticipates that the impact of these restrictions will diminish as adolescents grow older and become more independent. Furthermore, it posits that certain parenting practices, particularly those related to bedroom device access, will emerge as significant predictors of screen time trajectories. By incorporating passive measurement methods and examining these relationships longitudinally, this study offers a novel perspective on how parenting influences adolescent digital behavior. The findings are expected to inform targeted interventions that promote healthier digital habits among adolescents, especially during critical developmental periods when parental influence may wane. The ABCD data used in this report came from https://doi.org/10.82525/jy7n-g441. DOIs can be found at https://www.nbdc-datahub.org/abcd-release-6-0.

### Methods Participants

The ABCD Study is a prospective cohort study that enrolled 11,868 participants across 21 active study sites at baseline (Volkow et al., 2018). Participants complete annual lab assessments covering a wide range of measures, including screen usage questionnaires completed by youth and parents (Bagot et al., 2022). Passive smartphone use data were also collected through the ABCD-specific Effortless Assessment Research System (ABCD-EARS) application, which was installed on a subset of participants beginning at the year-3 visit (Wade et al., 2021).

The current analysis included participants who completed the year-3, year-4, and year-5 visits, as the Parent Screen Time Questionnaire (PSQ) was not administered prior to year 3. Data collection dates for this analysis ranged from January 2019 to January 2024, with participants ranging in age from 11 to 16 years across the three visits. Participants with missing visits were excluded, given the focus on longitudinal data and latent parenting profiles over time. The final analytical sample consisted of 8,096 participants, while 2,354 participants were excluded due to missing at least one visit. Comparisons between included and excluded participants are provided in Table 1.

**Table 1.** Comparison of Demographic and Household Characteristics Between Included and Excluded

|  | Included<br>(n=8096) | Excluded<br>(n=2354) | p value |
| --- | --- | --- | --- |
| **Interview Age (months (SD))** | 12.92<br>(0.64) | 13.08<br>(0.66) | <0.001 |
| **Sex** |  |  | 0.358 |
| Male | 4227<br>(52.2) | 1255 (53.3) |  |
| Female | 3869<br>(47.8) | 1099 (46.7) |  |
| **Household education** |  |  | <0.001 |
| < HS Diploma | 306 (3.8) | 131 (5.9) |  |
| HS Diploma/GED Post | 650 (8.1) | 234 (10.6) |  |
| Some College | 2432<br>(30.4) | 710 (32.2) |  |
| Bachelor | 1738<br>(21.8) | 460 (20.9) |  |
| Post Graduate Degree | 2861<br>(35.8) | 669 (30.4) |  |
| **Household income** |  |  | <0.001 |
| <50k | 1616<br>(20.2) | 544 (24.6) |  |
| ≥50K & <100K | 1939<br>(24.2) | 517 (23.4) |  |
| ≥100K | 3846<br>(48.0) | 898 (40.6) |  |
| Do not know | 362 (4.5) | 160 (7.2) |  |
| Refuse to Answer | 249 (3.1) | 95 (4.3) |  |
| **Parental Monitoring Survey (mean (SD))** | 4.38 (0.50) | 4.35 (0.52) | 0.007 |
| **Race/ethnicity** |  |  | 0.002 |
| White | 4544<br>(54.4) | 1164 (52.7) |  |
| Black | 1195<br>(14.3) | 265 (12.0) |  |
| Hispanic | 1645<br>(19.7) | 500 (22.6) |  |
| Asian | 159 (1.9) | 46 (2.1) |  |
| Other | 816 (9.8) | 235 (10.6) |  |
| **Self-Reported Total Screen Time**<br>**(hours/week)** | 4.56 (3.22) | 4.63 (3.33) | 0.331 |

**Note.** Included participants attended all required visits, while excluded participants missed at least one. P-values were estimated using a t-test for continuous variables and a chi-square test for categorical variables.

### Parent Screen Time Questionnaire

The Parent Screen Time Questionnaire (PSQ), introduced in year-3, consists of 14 parent-reported items (Figure 1) assessing media parenting practices, such as parental modeling, screen use during meals, screen time limits, and parental monitoring (Tang, Darlington, Ma, Haines, & Study, 2018). Items are rated on a 4-point Likert scale (1=Strongly Disagree, 2= Somewhat Disagree, 3=Somewhat Agree, 4=Strongly Agree). Refused responses (777) were coded as missing. Internal consistency, assessed via McDonald’s Omega, was excellent across visits (Year-3: ω = 0.92, Year-4: ω = 0.93, Year-5: ω = 0.93; Table S1).

**Figure 1.**
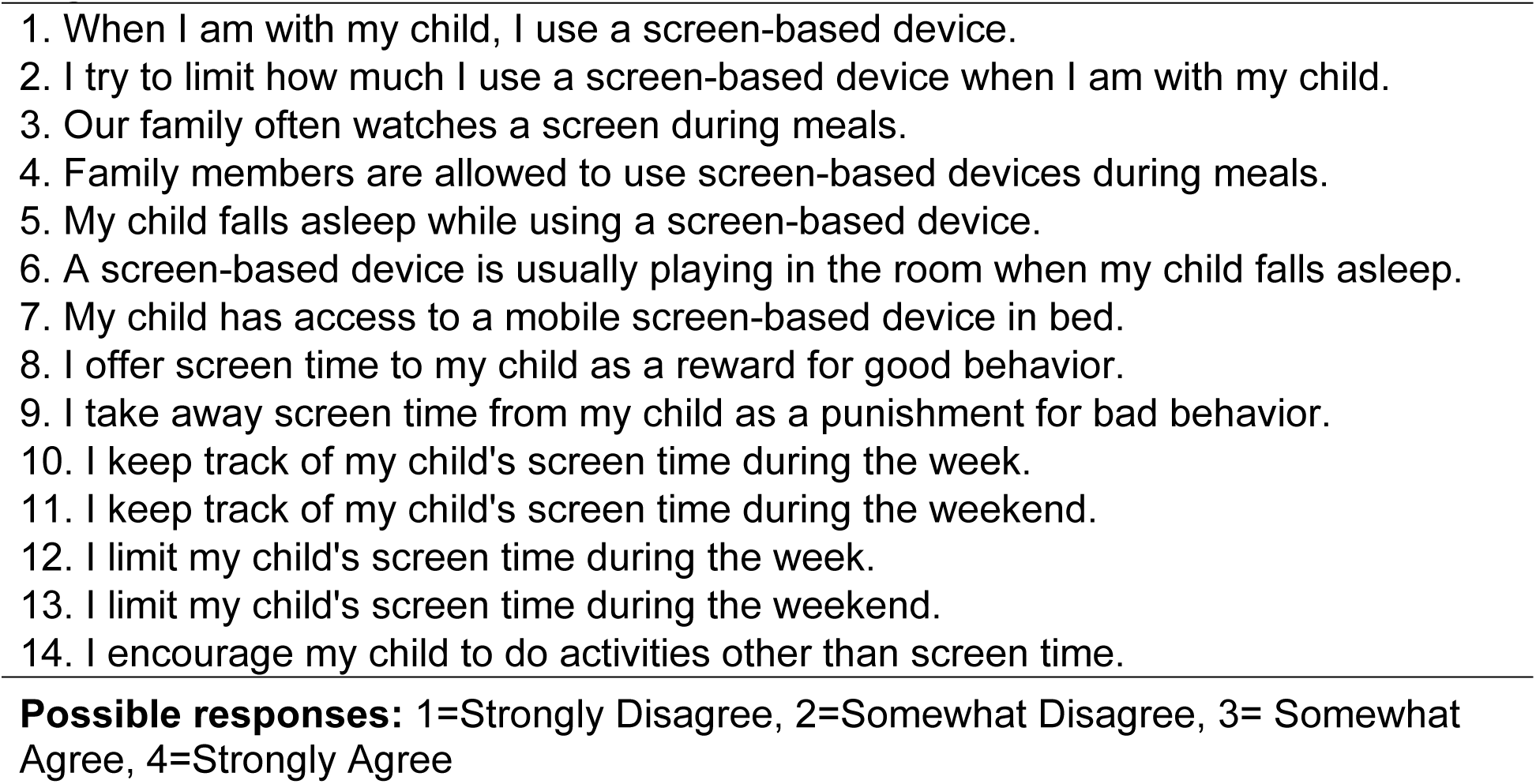
The Parent Screen Time Questionnaire

To maintain a consistent and intuitive interpretation, with higher scores reflecting greater restriction, items 1, 3, 4, 5, 6, 7, and 8 were reverse coded. This approach aligns with recommendations for latent transition analysis (LTA), ensuring clearer interpretation of class transitions (Weller, Bowen, & Faubert, 2020). During LTA, PSQ items were treated as categorical ordinal variables, which demonstrated superior model fit compared to treating items as continuous.

### Self-Reported Screen Time Measures

Self-reported total screen time was measured at years 3, 4, and 5, capturing time spent on a computer, phone, tablet, or other digital or mobile technology, as well as video games, excluding school-related work. Responses ranged from 0–23 hours (in 1-hour increments) and 0–45 minutes (in 15-minute increments). A summary score was calculated using the formula:

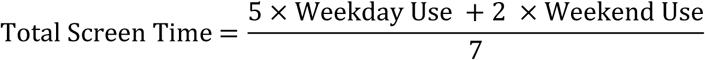

At years 4 and 5, a follow-up question assessed smartphone-specific screen time using the same formula. Due to a skip pattern, children without smartphones (9.8% at year 4) or those reporting zero total screen time were not asked about their smartphone use.

### Passive Assessment of Smartphone Use

Passive smartphone app use data were collected from a subset of participants using the ABCD-specific Effortless Assessment Research System (ABCD-EARS) application during the year-4 and year-5 visits (Wade et al., 2021). The app recorded daily screen use duration and app activity for Android devices only, as iOS devices do not permit passive app data collection. The analysis included 986 participants at year-4 and 885 at year-5, using the average daily screen use duration (nt_y_earsapp mins screenon nosys_mean) as the outcome variable.

### Passive Assessment of Keyboard Use

Keystroke data were collected via the ABCD-EARS app, using native keyboards on Android and third-party keyboards on iOS to circumvent operating system restrictions (Alexander et al., 2024). The analysis used the variable nt_y_earskey keyst_mean, representing the average number of daily keystrokes, as a proxy for smartphone engagement. This measure was particularly important because it allowed for the inclusion of data from iOS users, who represented the majority of device types in the ABCD-EARS study sample. Without this measure, no passive smartphone use data would have been available for iOS users. To account for device differences, models using keystroke data as the outcome included adjustments for the operating system. The final analytic sample included 2,846 participants at year 4 and 1,900 at year 5.

### Covariates

Covariates included age, sex, household income, parental education, study site, and youth-reported parental monitoring. Parental monitoring was assessed using the youth-reported Parental Monitoring Survey, a 5-item questionnaire rated on a 5-point Likert scale that evaluates parents’ active efforts to track their child’s whereabouts and activities (Karoly, Callahan, Schmiege, & Feldstein Ewing, 2016; Stattin & Kerr, 2000). A mean score was calculated, with higher scores indicating greater monitoring. Additional details about covariates are available on the ABCD Data Documentation website (docs.abcdstudy.org).

### Statistical Analysis

Latent transition analysis (LTA) was conducted in Mplus (version 8.1) to identify parenting profiles based on PSQ responses across years 3, 4, and 5 (Muthén & Muthén, 2017). This approach models changes in class membership over time (Collins & Lanza, 2009). Data preparation was performed using the MplusAutomation package in R (Hallquist & Wiley, 2018). Latent class models for each time point were fit using the 14 PSQ items as indicator variables to inform model selection. Model selection was guided by the following recommended fit indices: Constant Akaike Information Criterion (CAIC), sample-size adjusted Bayesian Information Criterion (BIC), entropy (>0.8), and posterior probabilities (Weller et al., 2020). A three-class model, representing Low Restriction, Moderate Restriction, and High Restriction groups was selected as the best compromise between parsimony and interpretability. Parameter restrictions, assuming measurement invariance across time, were imposed to ensure consistent class interpretation, improving stability and convergence (Collins & Lanza, 2009). LTA model fit indices and the fit indices with and without parameter restrictions are included in the supplementary materials (Table S2, Table S3, and Table S4).

Following LTA, class membership at each time point was exported to R for mixed effects analysis using the glmmTMB package (Brooks et al., 2017). Linear mixed-effects models were used to examine the relationships between parenting profiles and screen use outcomes, including self-reported total screen time (three time points), smartphone-specific screen time (two time points), passive assessment of smartphone use (two time points), and passive keystroke data (two time points). A random intercept for family ID and study site was included to account for the non-independence of observations within families and across sites (Dick et al., 2020). Separate models examined the interaction between age and class profile to assess how age influences the relationship between parenting restrictions and screen outcomes. Covariates included age, sex at birth, household income, parental education, and study visit. Beta coefficients with 95% confidence intervals were reported for the Moderate and High Restriction groups, using the Low Restriction group as the reference category.

Multicollinearity was assessed with the performance package in R by using variance inflation factors, which were all below 2.5 (Lüdecke, Ben-Shachar, Patil, Waggoner, & Makowski, 2021). To account for multiple comparisons, the Bonferroni correction was applied, setting a more stringent threshold for statistical significance. This approach was selected due to its strong control over Type I error, particularly in large-scale studies like ABCD, where extensive multiple testing increases the likelihood of false positives (VanderWeele & Mathur, 2019). All p-values reported are Bonferroni-adjusted for multiple comparisons. Replication code for all analyses is available on GitHub: https://github.com/Daniel-Adan-Lopez/ABCD_ParentProfiles.

### Machine Learning Methods

To complement these analyses, a gradient boosting decision tree model (XGBoost) was employed using the XGBoost package in Python (T. Chen, 2015; Van Rossum & Drake, 1995) to identify the most important parenting practices (PSQ items) contributing to self-reported youth screen time. The predictor set included all PSQ items and the visit variable to account for the longitudinal nature of the data. This approach was selected due to its ability to handle nonlinear relationships, interactions, and mitigate issues related to high correlations between PSQ items (Figure S1).

### Data Preprocessing and Splitting

The dataset was first filtered for completeness, with listwise deletion applied only to missing outcome values, allowing for the inclusion of observations with missing PSQ responses. This resulted in a final dataset of 24,198 observations across 8,095 unique participants. To account for within-family dependencies, participants were split by family ID, ensuring that siblings were assigned to the same dataset. This approach follows practices previously applied in the ABCD Study (Gray, Schvey, & Tanofsky-Kraff, 2020). The final 80/20 testing/training split resulted in 19,380 training observations from 6,484 participants and 4,818 testing observations from 1,611 participants.

### Model Training and Optimization

Hyperparameter optimization was performed via five-fold cross-validation and a randomized search strategy. The optimal model was selected based on the combination of hyperparameters that maximized the coefficient of determination (R²) and minimized root mean squared error (RMSE) during validation. The final model included 700 boosting iterations (n_estimators) with a maximum tree depth of 3, a learning rate (eta) of 0.01, and a gamma value (minimum loss reduction) of 0.1. Feature sampling was set to 0.9 (colsample_bytree), while row sampling (subsample) was 0.8, and the minimum child weight was 7.

### Feature Importance Analysis

Feature importance was assessed using Normalized Gain, which quantifies the relative contribution of each predictor to the model’s performance while ensuring that importance values sum to 1. Gain represents the average improvement in predictive accuracy when a feature is used in a decision split, with higher values indicating a greater contribution to reducing prediction error (T. Chen, 2015).

Normalization allows for direct comparison between predictors by rescaling their relative importance, ensuring interpretability across different features.

## Results

The characteristics of participants included (n=8359) and excluded due to having a missing time point (n=2354) showed no significant differences on the youth-reported Parental Monitoring Survey or on the youth-reported total screen time at the year-3 visit (Table 1). The included sample had an average age of approximately 13 years old and was skewed toward higher income and greater household education levels.

The distribution of individual PSQ items indicated an overall tendency toward more restrictive media parenting practices (Table S6). For instance, 70.4% of parents somewhat agreed or strongly agreed with taking away screen time as a form of punishment. Similarly, 65.6% of parents agreed or strongly agreed with keeping track of their child’s screen time during the week. Only 5% of responses disagreed with encouraging children to do activities other than screen time, and only 15.2% somewhat or strongly disagreed with limiting their own use of a screen-based device when around the child.

### Parenting Profiles Over Time

The distribution of parenting profiles from Year 3 to Year 5 revealed a significant shift toward less restrictive parenting practices regarding media use (Table 2). In Year 3, the Low Restriction profile accounted for 14.4% of participants (n=1164), increasing to 24.7% (n=1998) in Year 4 and 36.1% (n=2919) in Year 5. The High Restriction profile decreased from 46.9% (n=3796) in Year 3 to 35.2% (n=2849) in Year 4 and 25.3% (n=2044) in Year 5.

**Table 2.** Distribution of covariates by latent transitional class profile at year 3, year 4, and year

|  | Year 3 |  |  | Year 4 |  |  | Year 5 |  |  |
| --- | --- | --- | --- | --- | --- | --- | --- | --- | --- |
|  | Low<br>Restriction<br>(n=1164) | Moderate<br>Restriction<br>(n=3129) | High<br>Restriction<br>(n=3796) | Low<br>Restriction<br>(n=1998) | Moderate<br>Restriction<br>(n=3242) | High<br>Restriction<br>(n=2849) | Low<br>Restriction<br>(n=2919) | Moderate<br>Restriction<br>(n=3126) | High<br>Restriction<br>(n=2044) |
| **Interview Age (years(SD))** | 13.03 (0.64) | 12.96<br>(0.64) | 12.86<br>(0.64) | 14.25<br>(0.71) | 14.15<br>(0.70) | 14.02<br>(0.68) | 15.15<br>(0.67) | 15.07<br>(0.66) | 14.95<br>(0.66) |
| **Sex at birth** |  |  |  |  |  |  |  |  |  |
| Male | 504 (43.3) | 1588 (50.8) | 2133 (56.2) | 889 (44.5) | 1697 (52.3) | 1639 (57.5) | 1380 (47.3) | 1655 (52.9) | 1190 (58.2) |
| Female | 660 (56.7) | 1541 (49.2) | 1663 (43.8) | 1109 (55.5) | 1545 (47.7) | 1210 (42.5) | 1539 (52.7) | 1471 (47.1) | 854 (41.8) |
| **Household education** |  |  |  |  |  |  |  |  |  |
| < HS Diploma | 64 (5.6) | 86 (2.8) | 156 (4.1) | 91 (4.6) | 81 (2.5) | 124 (4.4) | 129 (4.5) | 80 (2.6) | 109 (5.5) |
| HS Diploma/GED Post | 137 (12.0) | 239 (7.8) | 273 (7.2) | 192 (9.7) | 276 (8.7) | 206 (7.3) | 252 (8.8) | 276 (9.0) | 154 (7.8) |
| Some College | 413 (36.3) | 1064 (34.5) | 955 (25.4) | 659 (33.4) | 1015 (31.8) | 666 (23.7) | 878 (30.5) | 917 (30.0) | 466 (23.5) |
| Bachelor | 218 (19.1) | 702 (22.8) | 818 (21.7) | 413 (20.9) | 719 (22.6) | 620 (22.0) | 644 (22.4) | 684 (22.4) | 431 (21.7) |
| Post Graduate Degree | 307 (27.0) | 990 (32.1) | 1564 (41.5) | 618 (31.3) | 1097 (34.4) | 1196 (42.5) | 974 (33.9) | 1101 (36.0) | 825 (41.6) |
| **Household income** |  |  |  |  |  |  |  |  |  |
| <50k | 295 (25.8) | 623 (20.2) | 697 (18.5) | 411 (20.8) | 571 (17.9) | 479 (17.0) | 513 (17.8) | 530 (17.3) | 328 (16.5) |
| ≥50K & <100K | 306 (26.7) | 802 (26.0) | 831 (22.0) | 451 (22.8) | 783 (24.5) | 542 (19.3) | 649 (22.5) | 651 (21.2) | 344 (17.3) |
| ≥100K | 446 (39.0) | 1450 (46.9) | 1950 (51.6) | 939 (47.5) | 1606 (50.3) | 1555 (55.3) | 1518 (52.6) | 1650 (53.8) | 1137 (57.2) |
| Do not know | 54 (4.7) | 119 (3.9) | 189 (5.0) | 86 (4.4) | 135 (4.2) | 150 (5.3) | 108 (3.7) | 123 (4.0) | 99 (5.0) |
| Refuse to Answer | 44 (3.8) | 95 (3.1) | 110 (2.9) | 89 (4.5) | 96 (3.0) | 88 (3.1) | 96 (3.3) | 112 (3.7) | 81 (4.1) |
| **Parental Monitoring (mean (SD))** | 4.31 (0.52) | 4.34 (0.50) | 4.45 (0.48) | 4.35 (0.49) | 4.37 (0.47) | 4.48 (0.45) | 4.36 (0.52) | 4.38 (0.49) | 4.49 (0.47) |
| **Self-reported total screen time, mean (SD) hours** | 5.55 (3.55) | 5.12 (3.29) | 3.80 (2.86) | 5.60 (3.47) | 5.17 (3.15) | 3.99 (2.83) | 5.19 (3.04) | 4.81 (2.83) | 3.93 (2.65) |

Note. Frequencies and percentages are presented unless otherwise indicated. Percentages are the proportions for each class profile at each time point. SD = Standard Deviation.

Significant differences were observed by sex, with males being more frequently assigned to the High Restriction group across all years compared to females. Conversely, females were more likely to belong to the Low Restriction group across Years 3 through 5.

High Restriction parenting was more prevalent in parents with postgraduate degrees (e.g., 41.5% in Year 3) and in households with incomes ≥$100,000 (e.g., 51.6% in Year 3). These patterns remained consistent over the three years.

Parental Monitoring scores were highest in the High Restriction group and increased slightly over time, from 4.45 (SD = 0.48) in Year 3 to 4.49 (SD = 0.47) in Year 5. The Low Restriction group consistently reported lower parental monitoring scores.

### Transition Probabilities

Parenting profiles remained relatively stable across the three time points (Table S5), with 21.5% of parents consistently in the High Restriction group, 17.8% in the Moderate Restriction group, and 10% in the Low Restriction group. Downward shifts in restriction levels were more common than upward shifts, suggesting that parents tended to relax restrictions as children aged.

The most common transition involved parents moving from Moderate Restriction at Years 3 and 4 to Low Restriction at Year 5 (8.6%). In contrast, upward transitions were rare, with only 0.03% shifting from Low → Moderate → High over the three years. Similarly, shifts from Low → Low → High were uncommon (0.1%). These patterns highlight a general trend of decreasing digital parenting restrictions over time, with few parents adopting stricter practices after initially adopting more permissive ones.

Final class counts and proportions are presented in Table S2. Conditional probabilities associated with these transitions are available in Table S5.

### Self-Reported Screen Time

On average, youth reported 4.74 hours of screen time per day across Years 3 through 5, with no significant difference between males (4.75 hours/day) and females (4.72 hours/day). Youth averaged 4.06 and 6.43 hours per day on weekdays and weekends, respectively.

As shown in Table 3, the High Restriction group reported about 56 fewer minutes of screen time per day compared to the Low Restriction group (95% CI: -49, -62.7, p < 0.001). The Moderate Restriction group reported 11.5 fewer minutes per day compared to the Low Restriction group (95% CI: -5.5, -17.5, p = 0.001).

**Table 3.**
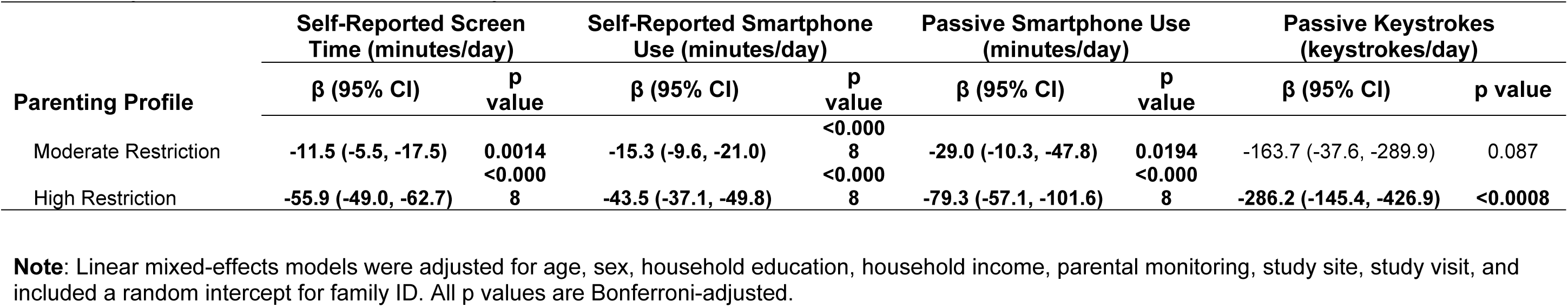
Longitudinal Associations Between Parenting Profiles and Media Use Outcomes (Reference = Low Restriction)

A significant interaction was observed between age and the High Restriction profile (Figure 2), indicating that as children grew older, those in the High Restriction group increased their screen time at a higher rate than those in the Low Restriction group (β = 13 minutes/day, 95% CI: 7.9, 18.2, p < 0.001) (Table 4). The Moderate Restriction profile did not significantly affect screen time trajectories.

**Figure 2.**
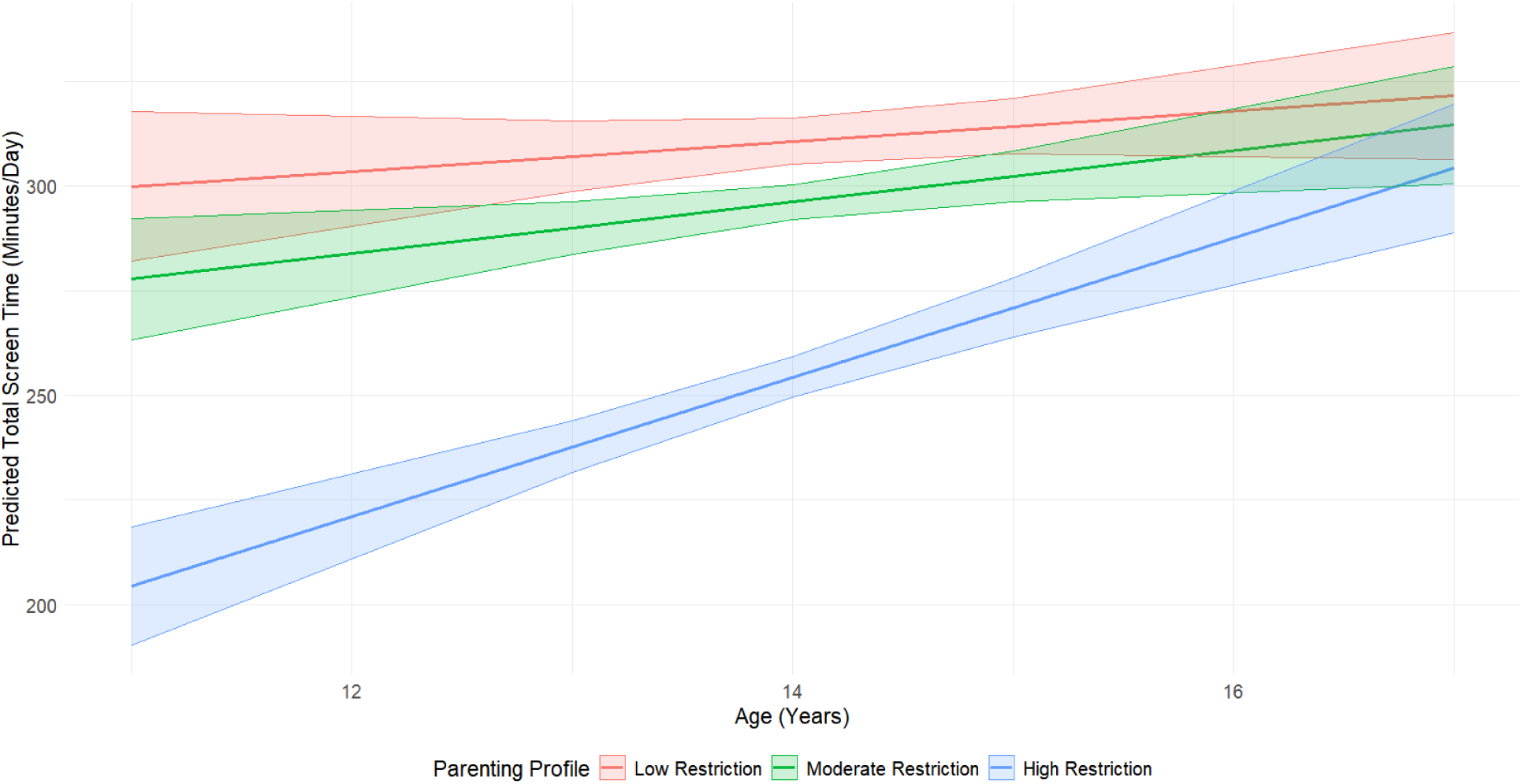
Interaction Effect of Parenting Profiles and Age on Predicted Self-Reported Screen Time. Predicted self-reported total screen time (minutes/day) by age, stratified by parenting profile. The High Restriction group (blue) shows a steeper increase in screen time with age compared to the Moderate (green) and Low Restriction (red) groups. Shaded areas represent 95% confidence intervals.

**Table 4.** Interaction Effects of Parenting Profiles and Age on Media Use Outcomes

| Interaction Term | Self-Reported Screen Time<br>(minutes/day) | Self-Reported Smartphone<br>Use (minutes/day) | Passive Smartphone Use<br>(minutes/day) | Passive Keystrokes<br>(keystrokes/day) |
| --- | --- | --- | --- | --- |
| | $\beta$ (95% CI), p value | $\beta$ (95% CI), p value | $\beta$ (95% CI), p value | $\beta$ (95% CI), p value |
| Moderate Restriction × Age | 2.54 (-2.4, 7.5), p = 1.000 | 5.32 (-1.6, 12.3), p = 1.000 | 7.01 (-13.9, 28), p = 1.000 | 30.1 (-123.9, 184.1), p = 1.000 |
| High Restriction × Age | **13.03 (7.9, 18.2), p < 0.0008** | 7.62 (0.01, 15.2), p = 0.398 | 9.81 (-13.3, 32.9), p = 1.000 | 52.79 (-115.8, 221.4), p = 1.000 |

**Note:** Linear mixed-effects models were adjusted for age, sex, household education, household income, parental monitoring, study site, study visit, and included a random intercept for family ID. All p values are Bonferroni-adjusted. **Interaction terms represent the estimated change in media use per additional year of age for children in each parenting profile, compared to the low restriction group.**

**Table 5.** Relative Contribution of Predictors to Youth Self-Reported Screen Time (XGBoost Model)

| Feature Description | Normalized Gain |
| --- | --- |
| A screen-based device is usually playing in the room when my child falls asleep. | 0.2296 |
| My child falls asleep while using a screen-based device. | 0.2094 |
| My child has access to a mobile screen-based device in bed. | 0.058 |
| Family members are allowed to use screen-based devices during meals. | 0.073 |
| I limit my child's screen time during the weekend. | 0.1016 |
| Our family often watches a screen during meals. | 0.0601 |
| I limit my child's screen time during the week. | 0.0346 |
| I take away screen time from my child as a punishment for bad behavior. | 0.0455 |
| I try to limit how much I use a screen-based device when I am with my child. | 0.0439 |
| I encourage my child to do activities other than screen time. | 0.0335 |
| When I am with my child, I use a screen-based device. | 0.0293 |
| I offer screen time to my child as a reward for good behavior. | 0.0197 |
| I keep track of my child's screen time during the weekend. | 0.0163 |
| I keep track of my child's screen time during the week. | 0.0161 |

**Note.** Normalized feature importance values from the XGBoost model, indicating the relative contribution of each predictor to youth-reported screen time. Higher values reflect greater predictive importance.

### Self-Reported Smartphone Screen Time

Youth self-reported approximately 3 hours of smartphone screen time per day across years 4 and 5, with large differences between male reported (2.5 hours per day) and female reported (3.64 hours per day) durations.

As shown in Table 3, the mixed-effects model found that the High Restriction group averaged 43.5 fewer minutes of smartphone screen time per day compared to the Low Restriction group (95% CI: 37.1, 49.8 minutes, p value < 0.0008) (Table 3). The Moderate Restriction group averaged about 15.3 fewer minutes of smartphone time per day (95% CI: 9.6, 21 minutes, p value < 0.0008) when compared to the Low Restriction group.

Although a significant interaction between age and the High Restriction group was initially detected (β = 7.6, 95% CI: 0.01, 15.2), it did not survive Bonferroni correction (p = 0.398) (Table 4).

### Passive Android Smartphone Use

Among youth who participated in ABCD-EARS and owned an Android device, the average smartphone use was 5.3 hours per day across years 4 and 5. Differences between males and females were modest, with males averaging 5.1 hours per day and females 5.5 hours per day. Notably, smartphone use increased by approximately 31 minutes per day from year 4 (5.0 hours) to year 5 (5.52 hours) The High Restriction group averaged 79.3 fewer minutes per day of passive smartphone use compared to the Low Restriction group (p < 0.0008) (Table 3). The Moderate Restriction group averaged 29 fewer minutes per day (95% CI: -10.3, -47.8, p = 0.019).

No significant interactions were observed between parenting profile and age for passive smartphone use (Table 4).

### Keyboard Strokes

Keystroke data were collected from both Android and iOS devices. On average, Android users recorded 1,442 keystrokes per day, while iOS users averaged 1,114 keystrokes per day. Across both operating systems, females consistently had higher average daily keystroke counts than males. Female participants averaged 1,764 keystrokes per day on Android devices and 1,319 keystrokes per day on iOS devices, compared to males, who averaged 1,207 and 864 keystrokes per day on Android and iOS devices, respectively.

The High Restriction group recorded 286 fewer keystrokes per day compared to the Low Restriction group (95% CI: -145, -427, p < 0.0008) (Table 3). The Moderate Restriction group recorded 163.7 fewer keystrokes per day, but this difference was not significant after correction (95% CI: -37.6, -289.9, p = 0.087).

No significant interactions were found between parenting profile and age for keystroke activity (Table 4).

### Feature Importance Ranking

The XGBoost model explained 10.2% of the variance in youth-reported screen time (Test R² = 0.1022), with a root mean squared error (RMSE) of 176.9 minutes and a mean absolute error (MAE) of 128.4 minutes (Table S7).

The feature importance analysis revealed that having a screen-based device playing in the room when the child falls asleep was the strongest predictor of screen time (Normalized Gain = 0.23). In other words, this feature accounts for 23% of the model’s predictive capability. Other key predictors included falling asleep while using a screen-based device (0.21), having access to a mobile screen-based device in bed (0.06), limiting a child’s screentime during the weekend (0.10). Full feature importance rankings are provided in Table 4 and Figure S2.

## Discussion

This study explored the longitudinal relationship between digital parenting profiles and various screen use outcomes. To our knowledge, this is the first study to examine these relationships using both self-reported and passive smartphone use measures over multiple time points. Our findings revealed a consistent inverse association between parenting restriction and adolescent screen time. Specifically, higher levels of restriction were associated with reduced overall screen time, lower smartphone use, and decreased smartphone engagement across both self-reported and passively measured outcomes. However, the influence of these restrictions appeared to weaken as children aged, highlighting the need to consider developmental changes when evaluating long-term effectiveness.

Additionally, machine learning analyses identified device access in the bedroom as a key predictor of screen time. These results underscore the critical role of parenting in shaping children’s digital behaviors while highlighting potential limitations, such as the declining impact of restrictions over time, and opportunities for targeted interventions, particularly around the bedroom environment.

### Parenting Restrictions and Screen Use

Our results indicate that greater parenting restrictions regarding screen use are associated with lower overall screen times. However, this effect appears to diminish as children age, likely reflecting increasing autonomy, peer influence, and shifts in family dynamics. As children grow older, they may gain more unsupervised access to devices, making parental restrictions harder to enforce. This trend suggests that parenting strategies may need to adapt as children move into adolescence.

The existing literature generally supports the link between more restrictive digital parenting and reduced media use. For instance, a study in Spain found that primary school children without parental rules on computer use were significantly more likely to spend ≥2 hours per day playing computer games (Hoyos Cillero & Jago, 2011). Similarly, younger girls in families without TV-viewing rules had higher odds of watching ≥2 hours of TV daily (Hoyos Cillero & Jago, 2011). The study authors posited that older age in children was a risk factor for excessive consumption due to the greater availability of media devices in the bedroom.

Additionally, Brindova et al. (2014) found that adolescents aged 11 to 15 whose parents rarely or never enforced rules on TV or computer use had significantly higher odds of excessive consumption (OR = 1.76 and 1.50, respectively). The older age group had a nearly four-fold increased risk of excessive consumption relative to the younger age group (OR=3.91), possibly due to greater availability of electronic devices in the bedrooms of older children (Brindova et al., 2014). Interestingly, restrictions related to content type (e.g., which shows children could watch) were not associated with excessive consumption, suggesting that time-based restrictions may be more effective than content-based controls. In line with these findings, Pyper, Harrington, and Manson (2016) reported that enforcing rules around screen time doubled the odds of meeting recommended screen time guidelines (OR = 2.03, p < 0.0001) among Canadian children aged 1–17. Taken together, these studies and our results emphasize that parenting restrictions play a crucial role in managing children’s media use, though their effectiveness varies by age.

### Age-Related Dynamics in Parenting Restrictions

The diminishing impact of parenting restrictions as children age may be explained by a combination of increased media access, peer influences, and developmental changes. A meta-analysis examining parental practices and screen time found a weak, negative correlation between child age and the use of restrictive mediation strategies, with restrictive practices being most effective in younger children (Griffiths, Benrazavi, & Teimouri, 2016). Although the correlation was weak, it underscores a gradual decline in the effectiveness of restrictive practices as children age, suggesting that parents may need to adapt their strategies accordingly.

Building on these findings, Poulain, Meigen, Kiess, and Vogel (2023) reported that restrictive media practices were far less common among older children, attributing this decline to parents’ increasing difficulty in enforcing rules as children grow older. Another study found that technology-related parenting strategies were not effective in reducing screen time in an adolescent sample (mean age = 14.4), while they were effective in younger (mean age = 4.8) and middle childhood (mean age = 9.3) groups (Sanders et al., 2016). The authors stressed the importance of realistic and developmentally appropriate parenting strategies.

Collectively, these findings highlight both the strengths and limitations of parental restrictions in moderating digital media use. They also raise important questions about which specific parenting practices remain effective as children transition into adolescence.

### Access to Media Devices in the Bedroom

Our machine learning analyses highlighted access to a device in the bedroom as a key contributor to self-reported screen use. This finding aligns with prior research suggesting that bedroom media access can significantly impact children’s media consumption patterns and sleep health.

The connection between bedroom media access and excessive media consumption, as well as disrupted sleep patterns, has been well documented. For example, Gentile, Berch, Choo, Khoo, and Walsh (2017) found that the presence of media devices in the bedroom was associated with poorer school performance and higher levels of physical aggression in children. Although the study focused on television and gaming devices, it underscores the broader implications of bedroom media access on developmental outcomes. Additionally, Brindova et al. (2014) found that children with a computer in their bedroom had more than twice the odds of excessive computer use (OR=2.25, p value <0.001). A study conducted in Portugal similarly found significantly greater television, laptop, and tablet screen time among children aged 6–10 who had these devices in their bedrooms, with computer use significantly increased among girls (Rodrigues et al., 2021).

The relationship between bedroom media access and sleep is particularly noteworthy. An experimental study involving adolescents found that turning off mobile phones in the hour before bedtime led to an average of 21 additional minutes of sleep per night (Bartel, Scheeren, & Gradisar, 2019). However, the study also reported high refusal rates during recruitment, as adolescents resisted changes to their technology access. Similarly, a longitudinal study conducted in Germany with adolescents found that restrictive parenting methods were not significantly associated with reducing smartphone use in the bedroom (p=0.445), suggesting that such restrictions may lose effectiveness as children grow older (Karsay, Schmuck, Stevic, & Matthes, 2023). The findings from both studies underscore a critical challenge for future interventions: adolescents’ reluctance to modify established digital habits, especially those tied to social connections and independence.

### Strengths

The strengths of this study include a large, diverse sample of participants assessed across multiple time points, enabling the investigation of trajectories in digital parenting restrictions and their associations with adolescent screen use. A major strength is the integration of both self-reported measures and passive data collected via the ABCD-EARS (Effortless Assessment Research System) application. The consistency between these two data sources strengthens confidence in the validity of our findings.

Additionally, this study extended beyond general parenting profiles by exploring individual PSQ items. This analysis identified the bedroom environment as a key contributor to adolescent screen time. Such findings have practical implications, as they can inform more targeted interventions focused on modifying bedroom dynamics to reduce excessive screen use.

### Limitations

Several limitations warrant consideration. First, our analytic sample was reduced by including only participants with complete data across all time points. However, comparisons between included and excluded participants revealed no significant differences in self-reported screen, suggesting that selection bias was likely minimal. Second, while the use of Latent Transition Analysis (LTA) provided insights into changes in parenting profiles over time, it limited the ability to highlight the effects of specific parenting behaviors. We addressed this by conducting a machine learning analysis of individual PSQ items, which identified key predictors of adolescent screen use. Third, the screen time outcomes had varying sample sizes due to the ABCD Study’s design. Specifically, participation in ABCD-EARS was limited to a subset of the sample, and passive smartphone duration data were only available for Android users. Ongoing data collection in future study waves will be essential to better understand these trajectories. Finally, the influence of the COVID-19 pandemic on our findings remains unclear. Given that data collection spanned from January 2019 to January 2024, it is likely that the Year-3 and Year-4 data were affected by stay-at-home orders and associated shifts in digital parenting practices. These effects may partially explain observed shifts in parenting profiles. However, developmental changes, such as increased autonomy as children age, likely contributed as well. Our statistical models adjusted for study site, which may mitigate variance introduced by state-level differences in pandemic responses. Future research should further explore how pandemic-related shifts in parenting practices impact long-term screen use and related health outcomes, such as obesity.

### Directions for Future Research

Age-related changes raise critical considerations for tailoring digital parenting strategies that evolve alongside adolescents’ developmental needs. Although restrictive practices, such as those observed in the High Restriction group, are associated with reduced screen time, their impact appears to diminish as children grow older. This pattern likely reflects increased autonomy, peer influence, and shifting family dynamics during adolescence.

Moreover, intervention strategies may be less effective in older age groups. For instance, a meta-analysis found that treatment effects aimed at reducing screen time were most pronounced in children aged 12 years or younger (Jones et al., 2021). The authors suggested that, given the wide variety of media devices now available, highly tailored behavioral strategies (e.g., goal setting and family social support) may be more effective than one-size-fits-all approaches. However, this tailored approach may come at the expense of generalizability, underscoring the need for flexible yet scalable interventions.

Future research should also explore the impact of removing media devices from the bedroom, with a particular focus on incorporating adolescent perspectives to avoid potential strain on the parent-child relationship. Given our findings on the bedroom environment’s role in screen use, interventions might benefit from combining device removal with open discussions about the health risks associated with excessive screen time, including its impact on sleep and mental health.

## Conclusion

Our findings suggest that greater digital parenting restrictions are consistently associated with lower smartphone use and overall screen time among adolescents. However, this effect appears to diminish as children grow older. Importantly, the bedroom environment emerged as a major determinant of overall screen time. These insights highlight areas where parents can intervene to reduce excessive screen use.

Beyond individual households, these findings have implications for educators, healthcare providers, and policymakers seeking to promote healthy digital habits among youth. Targeted interventions that address the bedroom environment and consider age-specific developmental needs may offer a practical pathway. Identifying critical developmental periods where specific types of restrictions exert the greatest influence on these outcomes will be vital for designing age-appropriate and effective interventions.

### Ethics Statement

The Adolescent Brain Cognitive Development℠ (ABCD) Study received institutional review board approval from each participating site, and informed consent/assent procedures were completed by all participants and their caregivers prior to data collection. The current study involved only secondary analysis of fully de-identified data obtained from the National Institute of Mental Health (NIMH) Data Archive and was therefore determined to be not human subjects research by the Oregon Health & Science University (OHSU) Institutional Review Board.

### Data Availability Statement

All data used in this study are publicly available through the National Institute of Mental Health (NIMH) Data Archive as part of the Adolescent Brain Cognitive Development℠ (ABCD) Study. Access requires registration and completion of a data use certification. Data can be accessed at: https://nda.nih.gov/abcd. Code used for all analyses is available on GitHub: https://github.com/Daniel-Adan-Lopez/ABCD_ParentProfiles

### Funding Statement

The authors received no specific funding for this work.

Data used in the preparation of this article were obtained from the <u>Adolescent Brain Cognitive</u> <u>Development℠ (ABCD) Study</u>, held in the NIH Brain Development Cohorts Data Sharing Platform. This is a multisite, longitudinal study designed to recruit more than 10,000 children age 9–10 and follow them over 10 years into early adulthood.

The ABCD Study® is supported by the National Institutes of Health and additional federal partners under award numbers U01DA041048, U01DA050989, U01DA051016, U01DA041022, U01DA051018, U01DA051037, U01DA050987, U01DA041174, U01DA041106, U01DA041117, U01DA041028, U01DA041134, U01DA050988, U01DA051039, U01DA041156, U01DA041025, U01DA041120, U01DA051038, U01DA041148, U01DA041093, U01DA041089, U24DA041123, U24DA041147. A full list of supporters is available at Federal Partners – ABCD Study.

A list of participating sites and a complete listing of the study investigators can be found on the Complete ABCD Study Roster. ABCD Consortium investigators designed and implemented the study and/or provided data but did not necessarily participate in the analysis or writing of this report. This manuscript reflects the views of the authors and may not reflect the opinions or views of the NIH or ABCD Consortium investigators.

### Author Contributions

- **Conceptualization:** Daniel A. Lopez
- **Methodology:** Daniel A. Lopez
- **Formal Analysis:** Daniel A. Lopez
- **Writing – Original Draft:** Daniel A. Lopez
- **Writing – Review & Editing:** Daniel A. Lopez, Bonnie J. Nagel, Gloria Ruiz-Orozco, Arturo Lopez-Flores
- **Visualization:** Daniel A. Lopez
- **Project Administration:** Daniel A. Lopez

---

**Figure S1.**
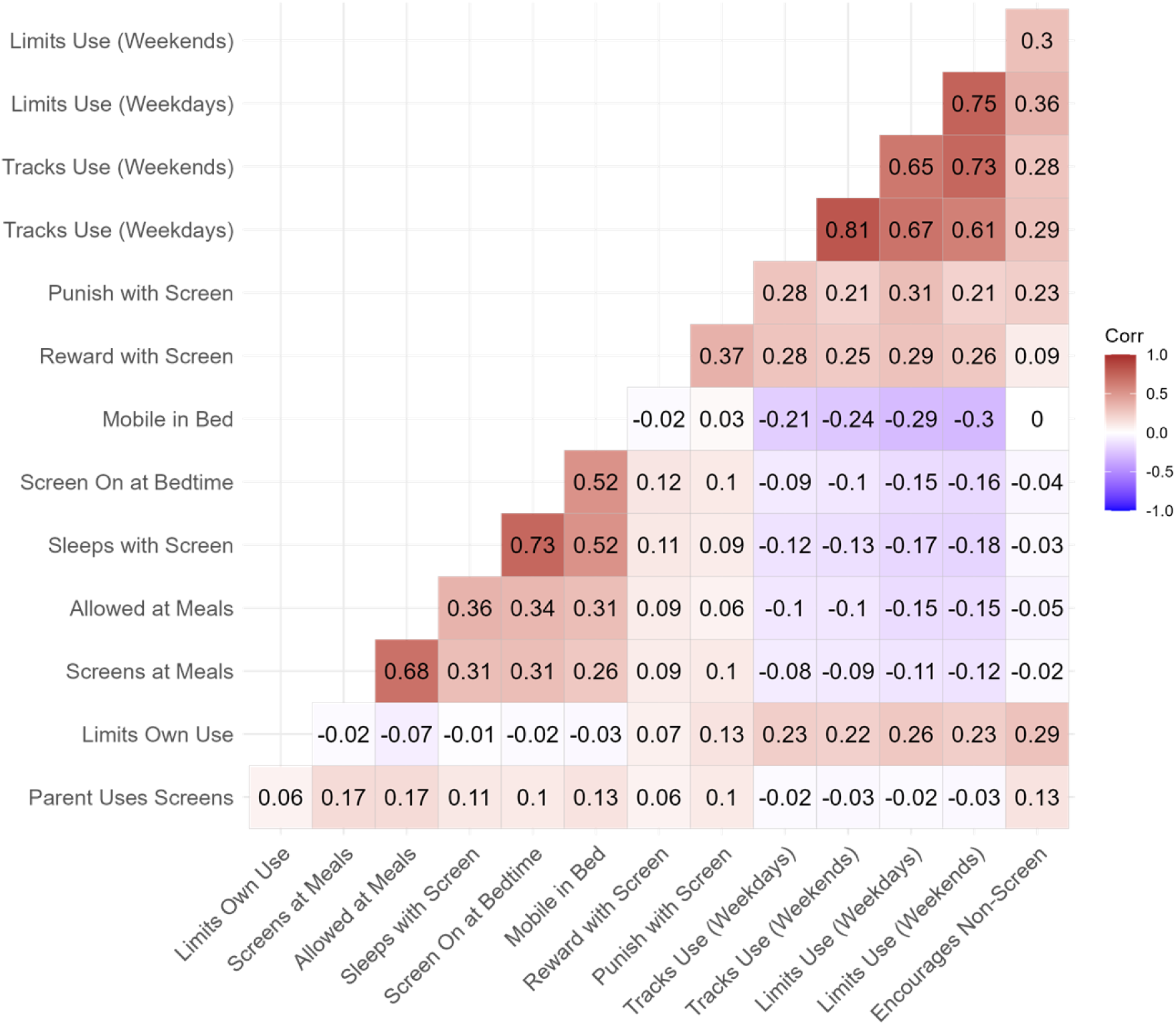
Correlation matrix of parental screen-related behaviors (PSQ items).

**Figure S2.**
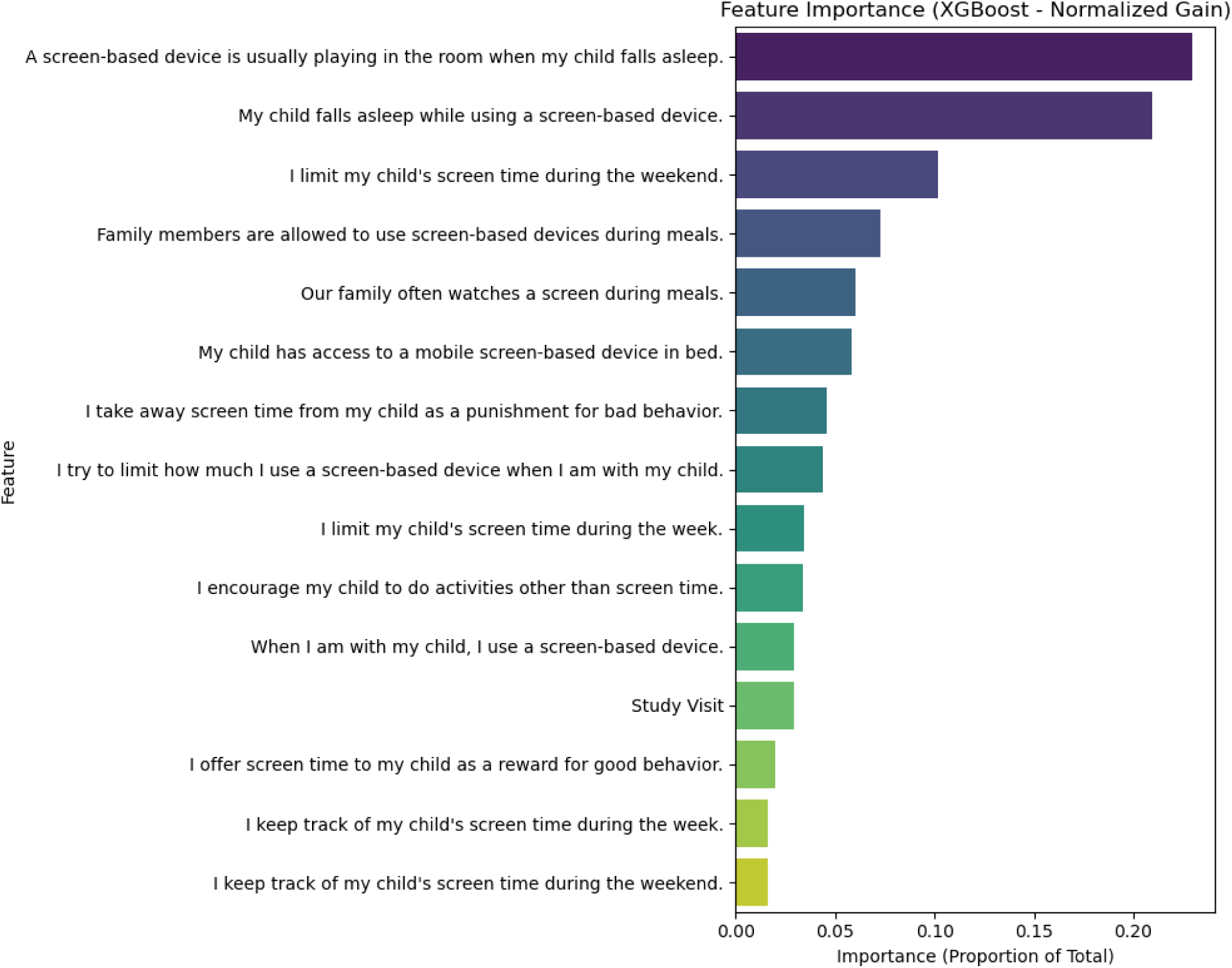
Feature Importance of Parenting Practices in Predicting Self-Reported Screen Time (XGBoost) XGBoost-derived feature importance (Gain) for parenting practices predicting self-reported total screen time. Higher Gain values indicate greater contribution to model predictions.

**Table S1.** Reliability Estimates (McDonald’s Omega) for PSQ Items Across Years 3, 4, and 5 McDonald’s Omega (ω)

| | McDonald's Omega ($\omega$) | | |
| --- | --- | --- | --- |
| | $\beta$ | SE | p-value |
| **Visit** |  |  |  |
| Year-3 | 0.918 | 0.001 | <0.001 |
| Year-4 | 0.929 | 0.001 | <0.001 |
| Year-5 | 0.933 | 0.001 | <0.001 |

**Table S2.** Final Class Counts and Proportions for Latent Class Trajectories Across Years 3, 4, and 5

| **Year 3** | **Year 4** | **Year 5** | **Count** | **Proportion (%)** |
| --- | --- | --- | --- | --- |
| **High** | **High** | **High** | **1,739.0** | **21.5%** |
| High | High | Moderate | 633.0 | 7.8% |
| High | High | Low | 230.0 | 2.8% |
| High | Moderate | High | 55.0 | 0.7% |
| High | Moderate | Moderate | 556.0 | 6.9% |
| High | Moderate | Low | 250.0 | 3.1% |
| High | Low | High | 30.0 | 0.4% |
| High | Low | Moderate | 67.0 | 0.8% |
| High | Low | Low | 236.0 | 2.9% |
| Moderate | High | High | 89.0 | 1.1% |
| Moderate | High | Moderate | 75.0 | 0.9% |
| Moderate | High | Low | 14.0 | 0.2% |
| Moderate | Moderate | High | 56.0 | 0.7% |
| **Moderate** | **Moderate** | **Moderate** | **1,443.0** | **17.8%** |
| Moderate | Moderate | Low | 693.0 | 8.6% |
| Moderate | Low | High | 22.0 | 0.3% |
| Moderate | Low | Moderate | 172.0 | 2.1% |
| Moderate | Low | Low | 565.0 | 7.0% |
| Low | High | High | 38.0 | 0.5% |
| Low | High | Moderate | 11.0 | 0.1% |
| Low | High | Low | 20.0 | 0.2% |
| Low | Moderate | High | 3.0 | 0.0% |
| Low | Moderate | Moderate | 80.0 | 1.0% |
| Low | Moderate | Low | 106.0 | 1.3% |
| Low | Low | High | 12.0 | 0.1% |
| Low | Low | Moderate | 89.0 | 1.1% |
| **Low** | **Low** | **Low** | **805.0** | **10.0%** |

Note. High, Moderate, and Low refer to restriction levels.

**Table S3.** Latent Transition Analysis Model Fit Indices for Year 3 to Year 5 (n=8096)

| Number of<br>Classes | LL | BIC | SABIC | CAIC | Entropy | Smallest off-<br>diagonal<br>ALCPP | Average latent class<br>posterior probabilities |
| --- | --- | --- | --- | --- | --- | --- | --- |
| 2 | -368100 | 737002 | 736719 | 736379 | 0.868 | 0.936 | 0.972, 0.936 |
| 3 | -352149 | 705557 | 705112 | 704578 | 0.876 | 0.919 | 0.949, 0.919, 0.933 |
| 4 | -343633 | 689030 | 688407 | 687658 | 0.886 | 0.923 | 0.923, 0.933, 0.944, 0.923 |
| 5 | -339385 | 681064 | 680254 | 679280 | 0.875 | 0.881 | 0.881, 0.889, 0.937, 0.91, 0.993 |
| 6 | -335460 | 673782 | 672772 | 671557 | 0.871 | 0.871 | 0.871, 0.894, 0.933, 0.904, 0.888, 0.900 |

Note. LL = Log-Likelihood, CAIC = Consistent Akaike Information Criterion, BIC = Bayesian Information Criterion, SABIC = Sample-Size Adjusted Bayesian Information Criterion, ALCPP = Average Latent Class Posterior Probability

**Table S4.** Fit Statistics and Class Enumeration for Latent Class Analysis by Time Point

| Model | # of<br>classes | LL | BIC | SABIC | CAIC | Entropy |
| --- | --- | --- | --- | --- | --- | --- |
| Year-3 visit (T1) (n=7954) | 1 | -<br>128006 | 256390 | 256257 | 256097 | NA |
|  | 2 | -<br>120389 | 241543 | 241273 | 240949 | 0.816 |
|  | 3 | -<br>116047 | 233245 | 232838 | 232351 | **0.882** |
|  | 4 | -<br>113355 | 228247 | 227703 | 227053 | 0.881 |
|  | 5 | -<br>112144 | 226211 | 225531 | 224717 | 0.864 |
|  | 6 | -<br>111228 | 224766 | 223949 | 222971 | 0.866 |
| Year-4 visit (T2) (n=7920) | 1 | -<br>131956 | 264289 | 264156 | 263996 | NA |
|  | 2 | -<br>123426 | 247615 | 247345 | 247022 | 0.856 |
|  | 3 | -<br>118242 | 237633 | 237226 | 236740 | 0.871 |
|  | 4 | -<br>115641 | 232817 | 232274 | 231624 | **0.889** |
|  | 5 | -<br>114447 | 230816 | 230136 | 229323 | 0.873 |
|  | 6 | -<br>113348 | 229004 | 228188 | 227211 | 0.871 |
| Year-5 visit (T3) (n=7891) | 1 | -<br>134229 | 268836 | 268703 | 268543 | NA |
|  | 2 | -<br>124599 | 249962 | 249691 | 249369 | 0.924 |
|  | 3 | -<br>120104 | 241357 | 240951 | 240465 | **0.873** |
|  | 4 | -<br>117998 | 237531 | 236988 | 236339 | 0.886 |
|  | 5 | -<br>116630 | 235181 | 234501 | 233689 | 0.879 |
|  | 6 | -<br>115501 | 233309 | 232492 | 231516 | 0.868 |

Note. LL = Log-Likelihood, CAIC = Consistent Akaike Information Criteria, BIC = Bayesian Information Criterion, SABIC = Sample-Size Adjusted Bayesian Information Criterion

**Table S5.**
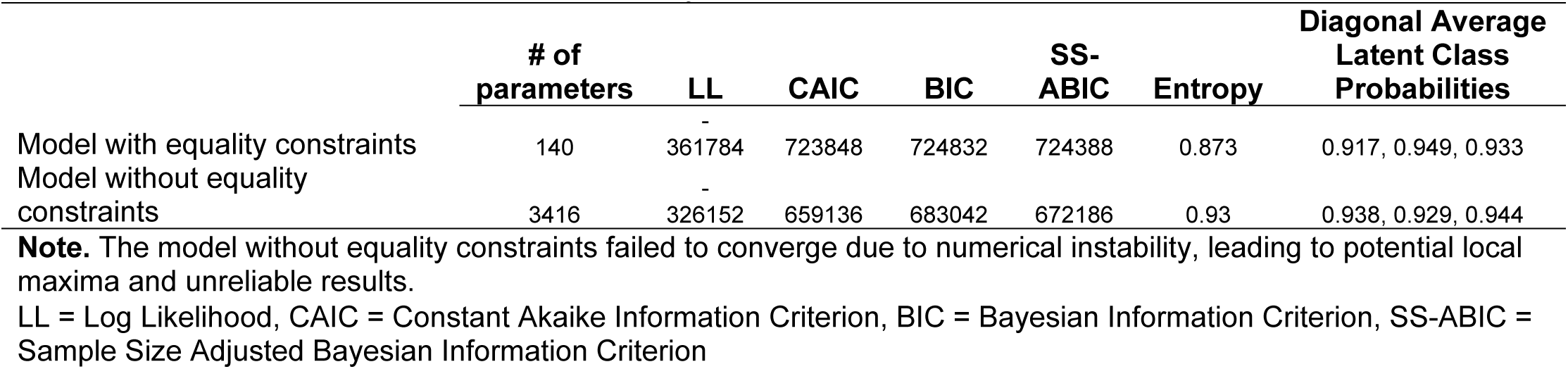
Fit Indices for Latent Transition Analysis Models With and Without Parameter Restrictions

**Table S6.** Frequency of Parent Responses to the ABCD Parent Screen Time Questionnaire (Years 3-5)

|  | **Strongly Disagree** | **Somewhat Disagree** | **Somewhat Agree** | **Strongly Agree** |
| --- | --- | --- | --- | --- |
| 1. When I am with my child, I use a screen-based device. | 2376 (10%) | 3557 (15%) | **11949 (50.5%)** | 5793 (24.5%) |
| 2. I try to limit how much I use a screen-based device when I am with my child | 1507 (6.4%) | 2090 (8.8%) | 9929 (42%) | **10136 (42.8%)** |
| 3. Our family often watches a screen during meals | **10689 (45.1%)** | 4462 (18.8%) | 5447 (23%) | 3117 (13.1%) |
| 4. Family members are allowed to use screen-based devices during meals | **12477 (52.6%)** | 4690 (19.8%) | 4568 (19.3%) | 1977 (8.3%) |
| 5. My child falls asleep while using a screen-based device | **13445 (56.7%)** | 3829 (16.2%) | 4527 (19.1%) | 1899 (8.0%) |
| 6. A screen-based device is usually playing in the room when my child falls asleep | **14351 (60.6%)** | 3167 (13.4%) | 3986 (16.8%) | 2190 (9.2%) |
| 7. My child has access to a mobile screen-based device in bed | **8596 (36.3%)** | 2291 (9.7%) | 5968 (25.2%) | 6829 (28.8%) |
| 8. I offer screen time to my child as a reward for good behavior | **13062 (46.1%)** | 3681 (16.0%) | 5013 (27.2%) | 1841 (10.6%) |
| 9. I take away screen time from my child as a punishment for bad behavior | 4881 (20.7%) | 2081 (8.8%) | 7129 (30.3%) | **9448 (40.1%)** |
| 10. I keep track of my child's screen time during the week | 5226 (22.1%) | 4693 (19.9%) | **8217 (34.8%)** | 5479 (23.2%) |
| 11. I keep track of my child's screen time during the weekend | 6174 (26.1%) | 5680 (24.0%) | **7453 (31.5%)** | 4336 (18.3%) |
| 12. I limit my child's screen time during the week. | 4489 (19.0%) | 3640 (15.4%) | 8311 (35.1%) | **7215 (30.5%)** |
| 13. I limit my child's screen time during the weekend. | 6013 (25.4%) | 6331 (26.8%) | **7235 (30.6%)** | 4080 (17.2%) |
| 14. I encourage my child to do activities other than screen time. | 797 (3.4%) | 387 (1.6%) | 3997 (17.0%) | **18277 (77.9%)** |

Note. Frequencies represent combined responses across years. Percentages represent proportion of response type by item.

**Table S7.** Conditional Probabilities of Class Membership at Year 5 Based on Year 3 and 4

| **Year 3 → Year 4** | **Probability of Low**<br>**Restriction at Year 5** | **Probability of**<br>**Moderate Restriction**<br>**at Year 5** | **Probability of**<br>**High Restriction**<br>**at Year 5** |
| --- | --- | --- | --- |
| Low → Low Restriction | **0.793** | 0.174 | 0.032 |
| Low → Moderate Restriction | 0.323 | **0.638** | 0.039 |
| Low → High Restriction | **0.101** | 0.26 | 0.639 |
| Moderate → Low Restriction | **0.793** | 0.174 | 0.032 |
| Moderate → Moderate<br>Restriction | 0.323 | **0.638** | 0.039 |
| Moderate → High Restriction | 0.101 | 0.26 | **0.639** |
| High → Low Restriction | **0.793** | 0.174 | 0.032 |
| High → Moderate Restriction | 0.323 | **0.638** | 0.039 |
| High → High Restriction | 0.101 | 0.26 | **0.639** |

**Note:** The table presents the conditional probabilities of class membership at Year 5 given transitions from Year 3 to Year 4. Cells with higher probabilities indicate the most likely class progression.

**Table S8.** Model Performance Metrics for Training and Test Sets

| **Metric** | **Training Set** | **Test Set** |
| --- | --- | --- |
| R<sup>2</sup> | 0.146 | 0.102 |
| Root Mean Squared Error | 172.7 | 176.9 |
| Mean Absolute Error | 124.5 | 128.4 |

**Note:** R<sup>2</sup> = The proportion of variance explained by the model

## References

1. Alexander, J. D., Linkersdörfer, J., Toda-Thorne, K., Sullivan, R. M., Cummins, K. M., Tomko, R. L., . . . Fuemmeler, B. F. (2024). Passively sensing smartphone use in teens with rates of use by sex and across operating systems. Scientific reports, 14(1), 17982.

2. Bagot, K., Tomko, R., Marshall, A., Hermann, J., Cummins, K., Ksinan, A., . . . Mason, M. (2022). Youth screen use in the ABCD® study. Developmental Cognitive Neuroscience, 57, 101150.

3. Bartel, K., Scheeren, R., & Gradisar, M. (2019). Altering adolescents’ pre-bedtime phone use to achieve better sleep health. Health communication, 34(4), 456–462.

4. Blum-Ross, A., & Livingstone, S. (2018). The trouble with “screen time” rules.

5. Brindova, D., Pavelka, J., Ševčikova, A., Žežula, I., van Dijk, J. P., Reijneveld, S. A., & Madarasova Geckova, A. (2014). How parents can affect excessive spending of time on screen-based activities. BMC public health, 14, 1–8.

6. Brooks, M. E., Kristensen, K., Van Benthem, K. J., Magnusson, A., Berg, C. W., Nielsen, A., . . . Bolker, B. M. (2017). glmmTMB balances speed and flexibility among packages for zero-inflated generalized linear mixed modeling. The R journal, 9(2), 378–400.

7. Chen, L., & Shi, J. (2019). Reducing harm from media: A meta-analysis of parental mediation. Journalism & Mass Communication Quarterly, 96(1), 173–193.

8. Chen, T. (2015). Xgboost: extreme gradient boosting. R package version 0.4-2, 1(4).

9. Collins, L. M., & Lanza, S. T. (2009). *Latent class and latent transition analysis: With applications in the social, behavioral, and health sciences* (Vol. 718): John Wiley & Sons.

10. Dick, A. S., Watts, A. L., Heeringa, S. G., Lopez, D. A., Bartsch, H., Fan, C. C., . . . Thompson, W. (2020). Meaningful Effects in the Adolescent Brain Cognitive Development Study. *BioRxiv*, 2020.2009.2001.276451. doi:10.1101/2020.09.01.276451

11. Gentile, D. A., Berch, O. N., Choo, H., Khoo, A., & Walsh, D. A. (2017). Bedroom media: One risk factor for development. Developmental Psychology, 53(12), 2340.

12. Gray, J. C., Schvey, N. A., & Tanofsky-Kraff, M. (2020). Demographic, psychological, behavioral, and cognitive correlates of BMI in youth: Findings from the Adolescent Brain Cognitive Development (ABCD) study. Psychological Medicine, 50(9), 1539–1547.

13. Griffiths, M., Benrazavi, R., & Teimouri, M. (2016). Parental mediation and adolescent screen time: A brief overview. Education and Health, 34(3), 70–73.

14. Hallquist, M. N., & Wiley, J. F. (2018). MplusAutomation: an R package for facilitating large-scale latent variable analyses in M plus. Structural equation modeling: a multidisciplinary journal, 25(4), 621–638.

15. Hoyos Cillero, I., & Jago, R. (2011). Sociodemographic and home environment predictors of screen viewing among Spanish school children. Journal of Public Health, 33(3), 392–402.

16. Jones, A., Armstrong, B., Weaver, R. G., Parker, H., von Klinggraeff, L., & Beets, M. (2021). Identifying effective intervention strategies to reduce children’s screen time: a systematic review and meta-analysis. International Journal of Behavioral Nutrition and Physical Activity, 18, 1–20.

17. Karoly, H. C., Callahan, T., Schmiege, S. J., & Feldstein Ewing, S. W. (2016). Evaluating the Hispanic paradox in the context of adolescent risky sexual behavior: The role of parent monitoring. Journal of pediatric psychology, 41(4), 429–440.

18. Karsay, K., Schmuck, D., Stevic, A., & Matthes, J. (2023). Sleeping with the smartphone: A panel study investigating parental mediation, adolescents’ tiredness, and physical well-being. Behaviour & Information Technology, 42(11), 1833–1844.

19. Lissak, G. (2018). Adverse physiological and psychological effects of screen time on children and adolescents: Literature review and case study. Environmental research, 164, 149–157.

20. Lüdecke, D., Ben-Shachar, M. S., Patil, I., Waggoner, P., & Makowski, D. (2021). performance: An R package for assessment, comparison and testing of statistical models. Journal of open source software, 6(60).

21. Muppalla, S. K., Vuppalapati, S., Pulliahgaru, A. R., Sreenivasulu, H., & kumar Muppalla, S. (2023). Effects of excessive screen time on child development: an updated review and strategies for management. Cureus, 15(6).

22. Muthén, B., & Muthén, L. (2017). Mplus. In Handbook of item response theory (pp. 507–518): Chapman and Hall/CRC.

23. Poulain, T., Meigen, C., Kiess, W., & Vogel, M. (2023). Media regulation strategies in parents of 4-to 16-year-old children and adolescents: a cross-sectional study. BMC public health, 23(1), 371.

24. Pyper, E., Harrington, D., & Manson, H. (2016). The impact of different types of parental support behaviours on child physical activity, healthy eating, and screen time: a cross-sectional study. BMC public health, 16, 1–15.

25. Rodrigues, D., Gama, A., Machado-Rodrigues, A. M., Nogueira, H., Rosado-Marques, V., Silva, M.-R. G., & Padez, C. (2021). Home vs. bedroom media devices: socioeconomic disparities and association with childhood screen-and sleep-time. Sleep Medicine, 83, 230–234.

26. Sanders, W., Parent, J., Forehand, R., Sullivan, A. D., & Jones, D. J. (2016). Parental perceptions of technology and technology-focused parenting: Associations with youth screen time. Journal of applied developmental psychology, 44, 28–38.

27. Stattin, H., & Kerr, M. (2000). Parental monitoring: A reinterpretation. Child development, 71(4), 1072–1085.

28. Tang, L., Darlington, G., Ma, D. W., Haines, J., & Study, G. F. H. (2018). Mothers’ and fathers’ media parenting practices associated with young children’s screen-time: A cross-sectional study. BMC obesity, 5, 1–10.

29. Van Rossum, G., & Drake, F. L. (1995). Python reference manual (Vol. 111): Centrum voor Wiskunde en Informatica Amsterdam.

30. VanderWeele, T. J., & Mathur, M. B. (2019). SOME DESIRABLE PROPERTIES OF THE BONFERRONI CORRECTION: IS THE BONFERRONI CORRECTION REALLY SO BAD? Am J Epidemiol, 188(3), 617–618. doi:10.1093/aje/kwy250

31. Volkow, N. D., Koob, G. F., Croyle, R. T., Bianchi, D. W., Gordon, J. A., Koroshetz, W. J., . . . Weiss, S. R. B. (2018). The conception of the ABCD study: From substance use to a broad NIH collaboration. Developmental Cognitive Neuroscience, 32, 4–7. doi:10.1016/j.dcn.2017.10.002

32. Wade, N. E., Ortigara, J. M., Sullivan, R. M., Tomko, R. L., Breslin, F. J., Baker, F. C., . . . Marshall, A. T. (2021). Passive sensing of preteens’ smartphone use: An Adolescent Brain Cognitive Development (ABCD) cohort substudy. JMIR mental health, 8(10), e29426.

33. Weller, B. E., Bowen, N. K., & Faubert, S. J. (2020). Latent class analysis: a guide to best practice. Journal of black psychology, 46(4), 287–311.

34. Zablotsky, B., Arockiaraj, B., Haile, G., & Ng, A. E. (2024). Daily Screen Time Among Teenagers: United States, July 2021–December 2023.

